# Exploring Needs and Priorities in Digital Health Management for Rare Disease Patients and their Caregivers: A Mixed-Methods Study

**DOI:** 10.64898/2026.01.28.26345095

**Authors:** Anita Burgun, Christina Khnaisser, Roxanne Dault, Jean-François Ethier

## Abstract

Rare diseases affect millions worldwide and are associated with long diagnostic delays, limited access to treatments, and substantial challenges in daily care and coordination. Digital health technologies, including mobile apps, telehealth, and data-sharing platforms, offer opportunities to improve care and quality of life for people living with rare diseases. As these tools rapidly expand, this study examines the needs, expectations, and conditions for successful adoption of patient-centered digital solutions among individuals living with rare diseases and their families.

Using a mixed-methods design, we surveyed 149 patients and caregivers, and conducted follow-up focus groups with 15 participants. Our findings highlight the essential role of digital tools in supporting people with rare diseases and their families. Key priorities include centralized health data, support for patient-generated data, and improved communication and information exchange with clinicians. Participants strongly emphasized the value of telehealth to reduce travel and simplify daily life, as well as patient-centered tools for diagnosis and emergency situations. Future digital solutions should integrate system-wide data, incorporate AI, and provide support during stressful situations, ultimately reducing patient burden despite persistent structural challenges. Respondents expressed strong interest in technologies that place patients at the center of care and improve coordination across providers.

Overall, our study identifies actionable targets for innovation and highlights technological, regulatory, and resource-related barriers that must be addressed to advance patient-centered digital solutions for rare diseases and guide future research and policy development.

**Author Summary:** People living with rare diseases often wait years for a diagnosis and struggle with complex, fragmented care. Digital health technologies could help address these challenges, but only if they are designed around patients’ real needs.

To better understand these needs, we surveyed 149 patients and caregivers in Quebec and held follow-up discussions with 15 participants. They emphasized the importance of centralized access to health information, better communication with clinicians, and tools that support patient-generated data. Telehealth was especially valued because it reduces travel and simplifies everyday life.

Our findings show that people with rare diseases want digital solutions that reduce their daily burden, improve coordination across providers, and support them during stressful moments such as emergency visits or the diagnostic process. This work provides practical guidance for designing patient-centered digital tools and highlights system-level barriers that must be addressed to ensure these innovations truly benefit the rare disease community.

## Introduction

Rare diseases are typically defined by a prevalence lower than 1 in 2,000 individuals [1] and consist of more than 7,000 identified types [2,3]. Worldwide, they impact over 300 million people [4], including approximately three million Canadians [3]. About 70% of these conditions begin in childhood [4], often resulting in a significant burden of intensive caregiving on families. Many rare diseases are associated with reduced life expectancy, as well as physical symptoms and disabilities that can limit daily activities, autonomy, and overall well-being [5,6].

Although rare diseases are highly heterogeneous and can affect multiple organs, their management presents common challenges. A primary issue is delivering an accurate and early diagnosis. Their diverse clinical presentations make it difficult for clinicians to identify and recall the symptoms, which can vary significantly within the same disease due to factors such as variable penetrance. In addition, their low prevalence means primary care physicians often encounter these diseases for the first time, leading to long delays between symptom onset and specialist referral. On average, rare disease patients wait five years for a diagnosis [7], and some remain undiagnosed even after referral. These “syndromes without a name” persist despite advances such as whole-genome sequencing (WGS). Initiatives like the Undiagnosed Disease Network in the US [8] and the Solve-RD project in the European Union [9] aim to close this gap, as faster and more accurate identification can significantly improve health outcomes and quality of life. Delayed diagnosis is often associated with higher costs, negative psychological impact, and greater social and functional consequences [10,11]. Moreover, an accurate diagnosis can help identify family members at risk, enabling genetic counseling and early screening.

Even after receiving a precise diagnosis, people with rare diseases continue to face multiple challenges, such as accessing quality care, coordinating hospital and outpatient services, maintaining autonomy, as well as achieving social, professional and civic integration. Above all, the greatest challenge remains access to appropriate treatments. This priority is reflected in a survey conducted by EURORDIS in 2018 involving 3,000 rare disease patients, where therapeutics ranked highest (8.4/10), followed by diagnosis (8.0), disease mechanisms (7.8), social, psychological and economic aspects (7.7), and assistance in daily life technologies (7.4) [12].

In 2025, acknowledging the challenges faced by rare disease patients and their families, the World Health Organization (WHO) designated rare diseases as a global health priority and urged countries to integrate this issue into their national strategies [13]. The WHO defined key components for global action, including (i) a framework to ensure equitable access to timely, cost-effective, and accurate diagnosis, evidence-based treatments, and adequate disease management; (ii) strategies for early and confirmed identification; and (iii) guidelines to improve access to healthcare services for rare disease patients. In addition to these recommendations, the WHO promotes the use of digital health technologies to improve access to specialists and treatments [13].

Digital health includes several technologies, such as mobile health applications, virtual care, wearable devices, and communication platforms such as social media. These technologies evolve rapidly. Clinician-patient communication has long been a cornerstone of medicine and patient-centered care, yet recent digital tools, mostly powered by large language models (LLMs), are increasingly integrated into these interactions [14]. They can facilitate patient data collection, analyze these data to provide recommendations, support documentation, and provide real-time responses to patient questions [15–17]. Telemedicine adoption has also drastically transformed healthcare delivery since the COVID-19 pandemic, with insurance programs worldwide expending coverage (e.g., U.S. Medicare and RAMQ in Québec [18,19]). While some patients are enthusiastic about digital tools, others remain skeptical, often due to prior negative experiences or a preference for human interaction [20]. Privacy concerns, particularly in rare diseases, should be explored to understand the extent to which individuals seek meaningful and transparent control over their health data [21,22]. This is especially relevant as patients with rare diseases may be more willing to share their data than other patients groups, given the challenges previously described [23].

While patients’ perception of digital tools is well documented in other domains [24,25], knowledge of rare disease patients’ expectations remains limited. Existing research has mainly focused on specific aspects of digital tool use in the rare disease community, particularly Internet use by parents of children living with a rare disease [26,27]. More recently, Chang et al. [28] explored perceptions of health digital tools among individuals with rare diseases and their caregivers, identifying three key roles: reducing the burden of disease management, fostering trust and community support, and ensuring access to reliable, personalized information.

Building on these insights, our study provides a complementary perspective through a mixed-methods design. Specifically, we aimed to: (i) describe interest and current use of digital health tools among individuals with rare diseases and their families; (ii) identify and prioritize patient-defined needs and preferences for patient-centered digital health solutions; and (iii) examine perceived challenges and conditions for successful implementation. By combining survey data with focus group insights, this study extends prior findings by offering a broader and more actionable understanding to inform the development and deployment of tailored digital health tools for the rare disease community.

## Methods

### Study design

To achieve our research objectives, we used a sequential mixed-methods design, starting with a survey followed by focus groups. The focus groups were designed to complement and enrich the survey by providing deeper context to the topics explored, rather than to serve as a means of validating the survey findings. Accordingly, individual survey responses were not matched with participants’ verbatim contributions during the focus groups.

### Recruitment

Members of the largest rare disease organization in Quebec (Canada), the *Regroupement Québécois des Maladies Orphelines* (RQMO), were targeted for this study [29]. Participants were eligible for the survey or the focus groups if they were 18 years old or older and if: (i) they had at least one diagnosis or rare disease or were in a diagnostic odyssey; or (ii) they were caregivers of a person with a rare disease (e.g., parent, partner).

The online survey was shared with the RQMO members via emails, newsletters, and social media posts on Facebook and LinkedIn. The survey link remained active for three months, during which several reminders were sent through these communication channels to help maximize the response rate. While survey responses remained confidential, participants interested in participating in the focus groups were invited to complete a separate form to provide their contact details. The survey was conducted between March and May 2024, and focus groups were held in April 2025.

### Data collection

#### Survey

The survey questionnaire was specifically developed by the authors for this study. Questions were designed to address the study objectives and included multiple-choice items and Likert scales responses. To ensure the relevance and clarity of the content, it was reviewed by two patient-partners from the RQMO. Additionally, an organization specializing in adult literacy (www.alphare.ca) assessed the language used to ensure accessibility for a broad audience. The survey was administered online in both French and English using the University of Sherbrooke LimeSurvey platform.

In the survey, respondents were first asked to assess their current use of digital tools and their interest in using them to access information, support the medical management of their condition, and assist with daily activities. They were then invited to identify the number one priority within these three domains. The survey also examined perceived challenges related to their development and implementation. At the end of the questionnaire, we included a series of sociodemographic and rare disease-related questions to help characterize the respondent sample.

The survey consisted of 27 core questions, along with 8 additional items related to sociodemographic and rare disease characteristics. It took approximately 20 minutes to complete. **S1** presents the survey questionnaire in English.

#### Focus groups

Focus groups consisted of survey respondents who had expressed interest in participating in this second phase of the study. Groups were formed based on participants’ availability and were separated into patients and caregivers to encourage discussions around shared experiences. Due to the diversity of rare diseases, we did not attempt to group participants according to their specific rare disease. This approach is also aligned with our objective to identify digital health needs, priorities and challenges that are common across the broader rare disease community.

Participants were required to sign an electronic consent form and complete a sociodemographic questionnaire before taking part in the discussion. The focus groups were conducted online using Microsoft Teams to enable participation of individuals located across the vast province of Quebec, including those in remote areas. This approach enriched the discussions, particularly given the relevance of digital tools for people living far from major urban centers. The focus groups were conducted in French, as all interested participants felt more comfortable to discuss in that language.

All focus groups were led by the same facilitator (RD), who has prior experience with this type of research design. A technical assistant (RM) supported each session by taking notes and managing logistical aspects. To capture both verbal and non-verbal cues, discussions were video, and audio recorded. A PowerPoint presentation was used to present the agenda, provide instructions, and guide participants through discussion topics and questions.

Participants were asked to keep their cameras and microphones open throughout the session to foster interaction and support group dynamics. When possible, they were encouraged to use the “raise hand” feature in Teams to indicate their intention to speak. The facilitator managed speaking turns to ensure fair and inclusive participation.

Each focus group followed the same structured discussion guide and lasted approximately 90 minutes. Groups were limited to a maximum of five participants, allowing meaningful exchanges and ensuring that everyone had the opportunity to answer and elaborate on all questions. A total of four focus groups, three with rare diseases patients and one with caregivers, were conducted in April 2025.

The qualitative phase was designed to enrich and contextualize the survey findings. To initiate the conversation and create a comfortable environment, participants were first asked whether they currently use digital tools to manage their rare disease. The discussion then explored their digital needs across key stages of their care journey: the diagnostic process, medical management following diagnosis, and day-to-day activities. Building on the survey findings, participants were sometimes asked specific questions about needs that had emerged as particularly significant. Finally, they were invited to identify what they considered the top priority for the development or implementation of digital health tools in Quebec. **S2** presents the discussion guide in English.

### Ethical approval

Ethics approval was obtained from the Educational and Social Sciences Research Ethics Committee of the University of Sherbrooke, Québec, Canada (approval number: 2023-4224). Informed consent was obtained online from all participants prior to their involvement in the study.

Participants’ contact information was stored in a secure online file accessible only by authorized members of the research team. Additionally, the qualitative data, such as the discussion transcripts, were stripped of all identifying information to preserve participant confidentiality.

### Data analysis

Qualitative findings were used to enrich the interpretation of the quantitative data and provide contextual depth, which was the primary purpose of adopting a mixed-methods design.

Survey findings were reported using descriptive statistical measures, such as proportions and mean values.

As the focus groups were audio and video recorded, the discussions were transcribed verbatim. To prevent participant re-identification, all identifying information, such as participants’ and rare disease names, was removed from the transcripts. Technical memos were inserted in brackets within the transcripts to capture nonverbal cues, hesitations, and explicit participant reactions. The transcripts were then imported into NVivo 12 qualitative analysis software for coding and interpretation. The coding tree combined both deductive and inductive approaches [30]. An initial high-level coding tree was developed in collaboration with the research team (AB, CK, JFE, RD), based on the themes addressed in the discussion guide. As the analysis progressed, new codes were added to reflect emerging themes and insights from the data. To ensure consistency in the analytical process, all focus groups were coded by the same member of the research team (RD). Regular team meetings were held to discuss emerging nodes and refine the coding tree as needed.

## Results

### Participant characteristics

A total of 149 individuals with rare diseases and caregivers completed an online survey conducted in Quebec (Canada) between March and May 2024. Subsequently, four focus groups were conducted in April 2025: three groups with patients and one with caregivers, each comprising three to five participants, for a total of 15 individuals.

Both patients and caregivers shared similar sociodemographic profiles (**Table 1**). Participants were predominantly female, generally under 55 years of age, and nearly half held a university degree. Almost all had received or cared for someone with an official rare disease diagnosis, mostly diagnosed within the past five years. Caregivers were primarily parents of minor children (0-17 years old). Almost all participants lived near a healthcare facility providing care for their rare disease, with most residing within one hour of travel and a significant proportion within two hours.

**Table 1.**
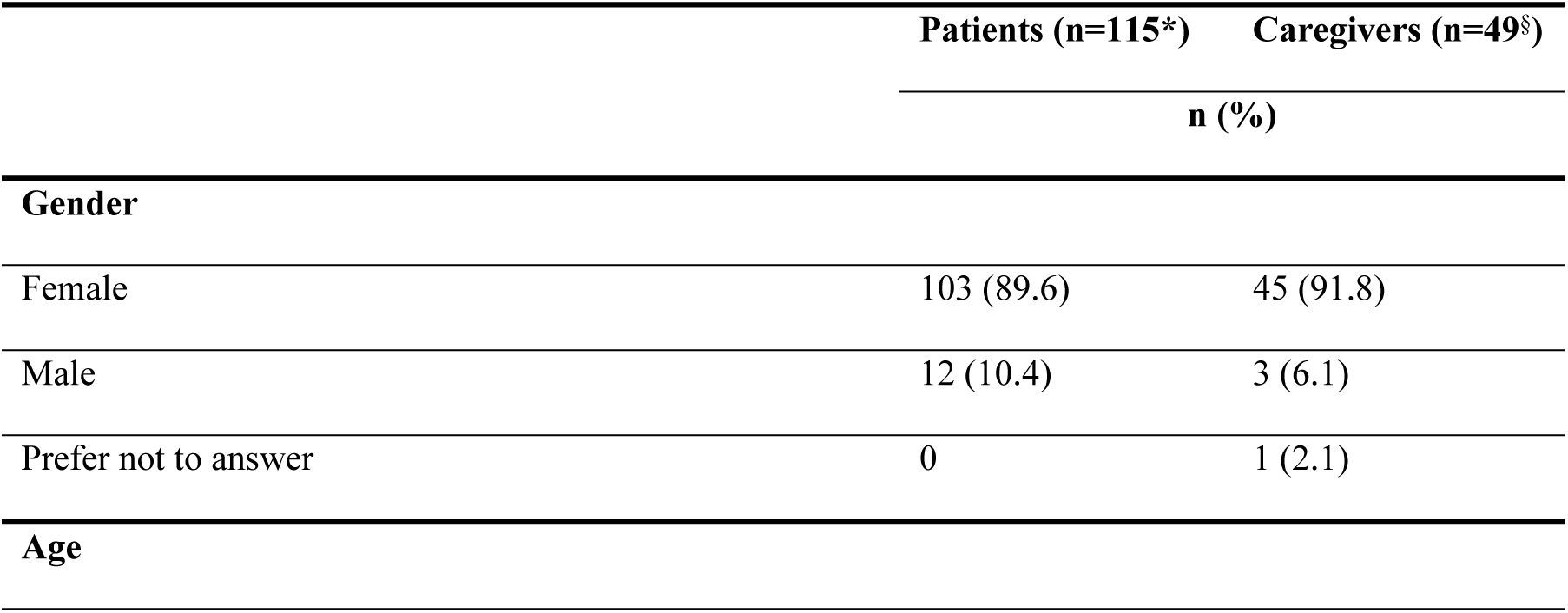
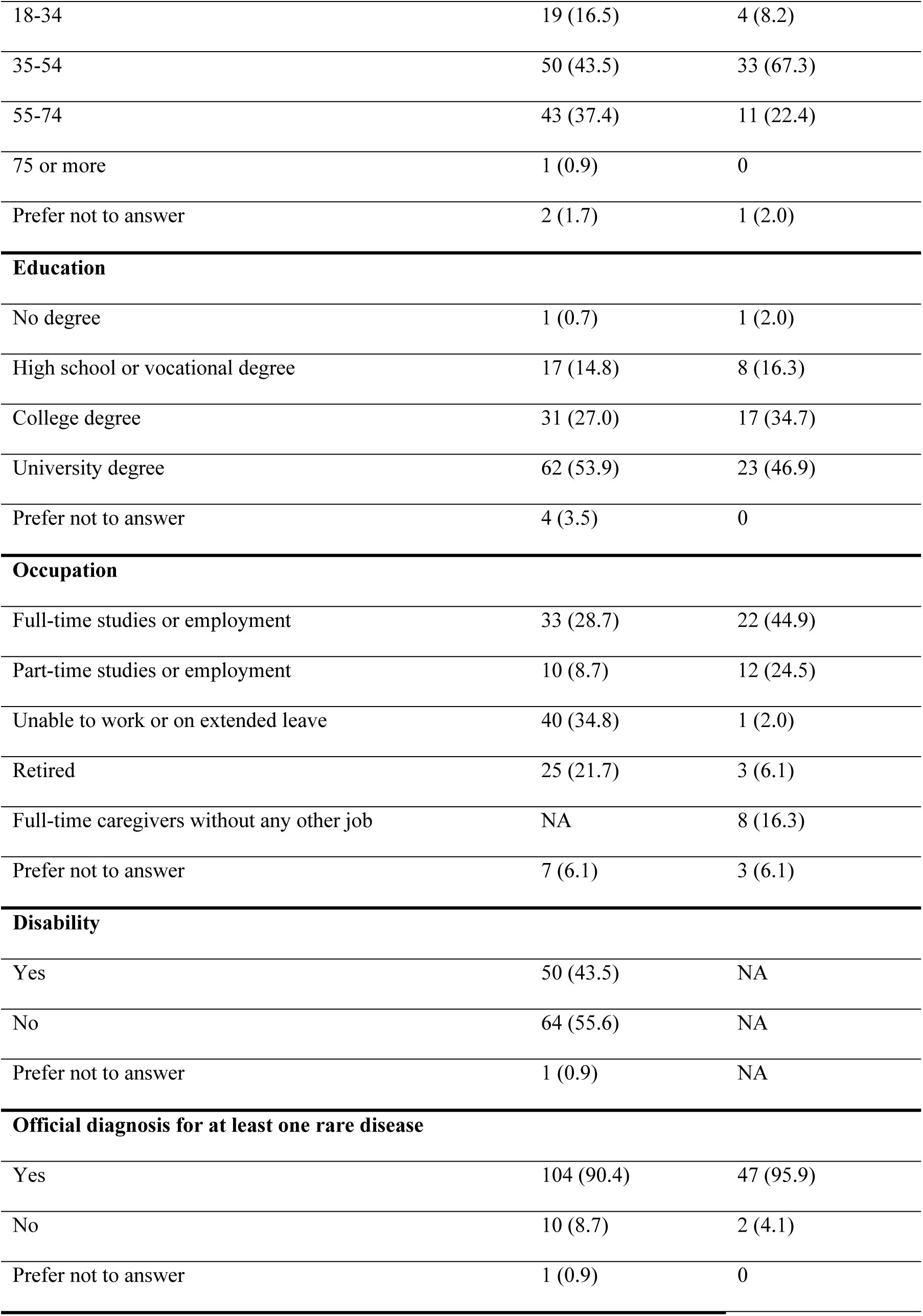
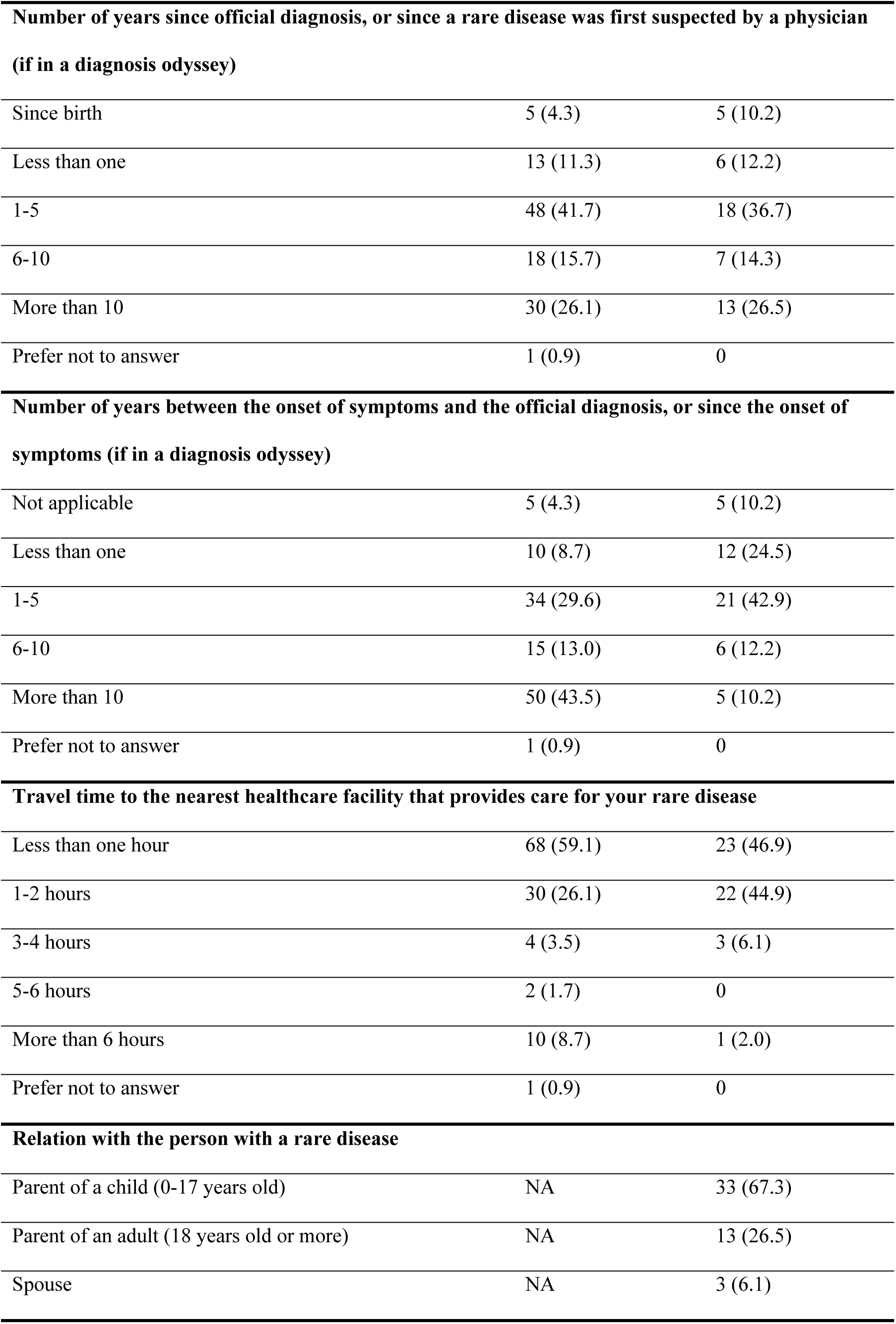
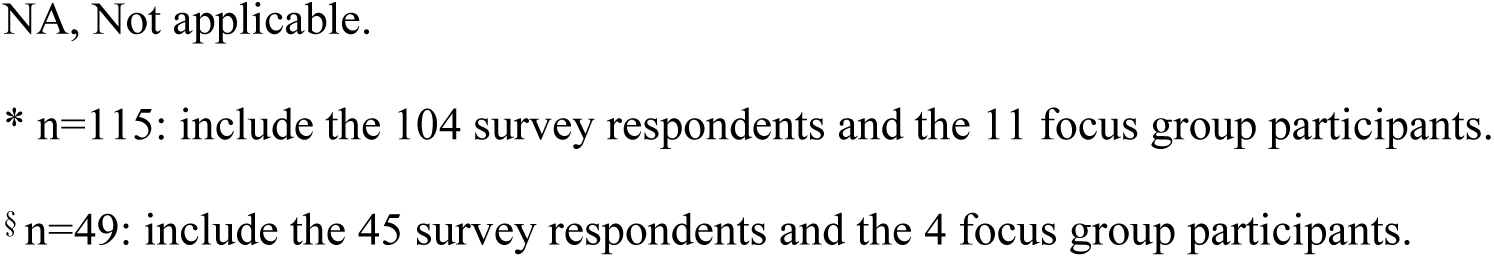
Characteristics of survey and focus group participants.

|  | Patients (n=115*) | Caregivers (n=49§) |
| --- | --- | --- |
|  | n (%) |  |
| Gender |  |  |
| Female | 103 (89.6) | 45 (91.8) |
| Male | 12 (10.4) | 3 (6.1) |
| Prefer not to answer | 0 | 1 (2.1) |
| Age |  |  |
| 18-34 | 19 (16.5) | 4 (8.2) |
| 35-54 | 50 (43.5) | 33 (67.3) |
| 55-74 | 43 (37.4) | 11 (22.4) |
| 75 or more | 1 (0.9) | 0 |
| Prefer not to answer | 2 (1.7) | 1 (2.0) |
| **Education** |  |  |
| No degree | 1 (0.7) | 1 (2.0) |
| High school or vocational degree | 17 (14.8) | 8 (16.3) |
| College degree | 31 (27.0) | 17 (34.7) |
| University degree | 62 (53.9) | 23 (46.9) |
| Prefer not to answer | 4 (3.5) | 0 |
| **Occupation** |  |  |
| Full-time studies or employment | 33 (28.7) | 22 (44.9) |
| Part-time studies or employment | 10 (8.7) | 12 (24.5) |
| Unable to work or on extended leave | 40 (34.8) | 1 (2.0) |
| Retired | 25 (21.7) | 3 (6.1) |
| Full-time caregivers without any other job | NA | 8 (16.3) |
| Prefer not to answer | 7 (6.1) | 3 (6.1) |
| **Disability** |  |  |
| Yes | 50 (43.5) | NA |
| No | 64 (55.6) | NA |
| Prefer not to answer | 1 (0.9) | NA |
| **Official diagnosis for at least one rare disease** |  |  |
| Yes | 104 (90.4) | 47 (95.9) |
| No | 10 (8.7) | 2 (4.1) |
| Prefer not to answer | 1 (0.9) | 0 |

| **Number of years since official diagnosis, or since a rare disease was first suspected by a physician**<br>**(if in a diagnosis odyssey)** |  |  |
| --- | --- | --- |
| Since birth | 5 (4.3) | 5 (10.2) |
| Less than one | 13 (11.3) | 6 (12.2) |
| 1-5 | 48 (41.7) | 18 (36.7) |
| 6-10 | 18 (15.7) | 7 (14.3) |
| More than 10 | 30 (26.1) | 13 (26.5) |
| Prefer not to answer | 1 (0.9) | 0 |
| **Number of years between the onset of symptoms and the official diagnosis, or since the onset of**<br>**symptoms (if in a diagnosis odyssey)** |  |  |
| Not applicable | 5 (4.3) | 5 (10.2) |
| Less than one | 10 (8.7) | 12 (24.5) |
| 1-5 | 34 (29.6) | 21 (42.9) |
| 6-10 | 15 (13.0) | 6 (12.2) |
| More than 10 | 50 (43.5) | 5 (10.2) |
| Prefer not to answer | 1 (0.9) | 0 |
| **Travel time to the nearest healthcare facility that provides care for your rare disease** |  |  |
| Less than one hour | 68 (59.1) | 23 (46.9) |
| 1-2 hours | 30 (26.1) | 22 (44.9) |
| 3-4 hours | 4 (3.5) | 3 (6.1) |
| 5-6 hours | 2 (1.7) | 0 |
| More than 6 hours | 10 (8.7) | 1 (2.0) |
| Prefer not to answer | 1 (0.9) | 0 |
| **Relation with the person with a rare disease** |  |  |
| Parent of a child (0-17 years old) | NA | 33 (67.3) |
| Parent of an adult (18 years old or more) | NA | 13 (26.5) |
| Spouse | NA | 3 (6.1) |

NA, Not applicable.
\* n=115: include the 104 survey respondents and the 11 focus group participants.
§ n=49: include the 45 survey respondents and the 4 focus group participants.

### Digital health needs for rare diseases

Results from rare disease patients and caregivers were combined, and no subgroup analysis was conducted as caregivers were asked to answer from the viewpoint of the individual they are caring for.

The survey participants showed interest in digital tools to solve a variety of problems (**Table 2**). The topics that generate the most interest include diagnosis and care trajectory management, which could both benefit from telemedicine tools. Better disease understanding is also important, and respondents indicated that information on websites would be sufficient. Moreover, around three out of four respondents consider that applications should be offered to support managing day-to-day activities and psychological well-being, and to support in finding social and education services. Finally, sharing experiences with other families through social media platforms is considered helpful by 87% of respondents.

**Table 2.** Survey respondents’ interest in using digital health tools to support rare disease management (n=149)

|  | Moderate to<br>high interest | Little to<br>no interest | Already<br>using | Digital solution<br>identified as the most<br>useful to meet this need |
| --- | --- | --- | --- | --- |
|  | n (%) |  |  |  |
| To find specialists doctors or specialized clinics for the rare disease | 129 (86.6) | 9 (6.0) | 11 (7.4) | App or platform that relies on personal data |
| To be informed about the progression of the medical condition related to the rare disease | 126 (84.6) | 11 (7.4) | 12 (8.0) | Telemedicine |
| To find clinical trials available for the rare disease and verify eligibility | 124 (83.2) | 20 (13.4) | 5 (3.4) | App or platform that relies on personal data |
| To monitor concerning symptoms or side effects after a treatment or after taking medication | 123 (82.6) | 20 (13.4) | 6 (4.0) | Telemedicine |
| To find social services and assistance programs (e.g., programs in mental health, social or financial aid, specialized education services, etc.) | 121 (81.2) | 26 (17.5) | 2 (1.3) | App or platform that relies on personal data |
| To better understand the rare disease or similar rare diseases | 117 (78.5) | 6 (4.0) | 26 (17.5) | Online information (Internet) |
| To maintain or improve psychological well-being | 110 (73.8) | 33 (22.2) | 6 (4.0) | Telemedicine |
| To talk with people who are going through similar experiences | 107 (71.8) | 19 (12.8) | 23 (15.4) | Forums or social media |
| To be informed about the risks of hereditary transmission of the rare disease | 103 (69.1) | 33 (22.1) | 13 (8.7) | Telemedicine |
| To receive support in managing day-to-day activities (e.g., adapting the diet, living environment, etc.) | 103 (69.1) | 41 (27.5) | 5 (3.4) | App or platform that relies on personal data |
| To manage health appointments | 102 (68.4) | 18 (12.1) | 29 (19.5) | App or platform that<br>relies on personal data |
| To suggest possible diagnoses based<br>on symptoms | 99 (66.4) | 32 (21.5) | 18 (12.1) | Telemedicine |
| To participate in online rehabilitation<br>programs | 94 (63.1) | 53 (35.6) | 2 (1.3) | Telemedicine |
| To manage medication | 86 (57.7) | 56 (37.6) | 7 (4.7) | App or platform that<br>relies on personal data |
| To monitor non-concerning<br>symptoms or side effects after a<br>treatment or after taking medication | 84 (56.4) | 61 (40.9) | 4 (2.7) | App or platform that<br>relies on personal data |

The functionalities for which at least two thirds of the participants showed little or no interest include drug management and monitoring of non-severe symptoms. Moreover, online rehabilitation programs were not a priority either, but not all rare diseases require such programs.

### Digital tools to support the diagnosis process

Most survey participants expressed moderate to high interest (78.5%) in using a digital tool to support the diagnosis process (**Table 2**). This highlights a significant gap in achieving timely diagnosis, as most participants had already received a diagnosis yet still expressed a strong desire for this kind of tool. This need was strongly emphasized during the focus groups. Several participants described the diagnostic process as “looking for a needle in a haystack.” Repeated testing often delays diagnosis, allowing the disease to progress and worsen:

> “*Sometimes they have to do the same test several times to reach a diagnosis. Patients often lose skills because the diagnosis takes so long. That clogs up the system even more*.” (P13)

One participant shared that they created an Excel spreadsheet to document all relevant information about their condition: symptoms, medications, treatments, and medical consultations, to facilitate and accelerate the diagnostic process:

> *“Before I got my diagnosis, I created an Excel spreadsheet with all my symptoms, my medication, what did the family doctor do, what helped the symptoms, etc. Instead of starting from scratch, they [doctors in Mexico, where he got his diagnosis] did all the preliminary work before I even went there. If there was a system like this [digital tool] here in Quebec or in Canada, it would be so easy.”* (P7)

Participants highlighted the value of a digital tool that could support the exploration of potential diagnoses. Digital tools also have the potential to help patients articulate what they feel and share it effectively with their medical team, while providing validation that what they experience is real. They can enable patient-reported signs and symptoms, allowing users to generate possible diagnostic leads. This is especially relevant in the era of self-quantified devices enabling comprehensive measurements during activities of daily living and sleep periods:

> *“If there was a search engine… My son has a hump on his back. Well, what genetic diseases can cause a hump on the back? Entering specific characteristics or clinical symptoms would have helped. You can search by keywords. I think that’s what’s missing. It would help a lot.”* (P14)

> *“I think symptom tracking could help get a diagnosis. I had my handwritten sheets. It would certainly have been easier. Then, you might see the impact of certain things. Sometimes we have symptoms, but we don’t know how to name them properly [other participants agree].”* (P10)

Interestingly, some participants expressed huge expectations regarding patient-centric tools, subsequently putting the patient (and not the physician) as the main actor in the diagnostic process:

> “*A Quebec tool that you could bring to your doctor and say, ‘I looked this up, what do you think?’ We are the patients; we are the ones who know the most about our condition.*” (P2).

However, the participants emphasized that for such a digital tool to be qualified as trustworthy and adopted by patients, it should be certified by national agencies.

### Digital tools to support disease management

Survey respondents expressed moderate to high interest in digital tools designed to support various aspects of their medical care (**Table 2**). These included five main topics: (i) better understanding of their rare disease (78.5%), including its progression (84.6%), (ii) finding specialists or specialized clinics (86.6%), (iii) managing healthcare trajectories (68.4%), (iv) monitoring adverse effects of treatments or medications, whether severe (82.6%) or not (56.4%); and (v) accessing research related to their rare disease and identifying available clinical trials (83.2%). These needs were also echoed in the focus groups.

#### (i) Better understanding of their rare disease, including its progression

In the focus groups, several participants reported lacking access to reliable tools for answering questions or verifying information about their condition, particularly regarding symptoms and disease progression over time. This is related both to the lack of knowledge about certain diseases and to the absence of tools tailored for patients, even when information does exist. They noted that existing tools are often difficult to use, not available in their language, and that the information provided is not made accessible to a general audience, limiting both usability and understanding. Therefore, they are looking for digital tools that are user-friendly and multilingual, can adapt to different levels of health literacy and provide trustworthy information:

> *“Orphanet is great, but at the same time, it’s not very user-friendly when it comes to searching. And even then, you have to know what you are looking for.”* (P14)

> *“I feel there’s a lot of misinformation out there. So, we need to be careful. Having a good platform for all this could only be beneficial.”* (P2)

> *“I searched a lot for simplified information about my disease, even just about the symptoms. I use the web a lot to try to find information.”* (P11)

Additionally, several participants expressed difficulties in being taken seriously when reporting unfamiliar symptoms to some health professionals and even in obtaining general information about their condition. Many rare disease patients consider that only family members, a handful of caregivers, and those suffering from a rare illness are aware of what they are going through, which makes them look for digitally mediated support to get more information:

> *“I have been saying for five years that I need to see a neuropsychologist, but because there is so little information available, doctors tell me, ‘No, your disease doesn’t cause cognitive problems.’ But on Facebook, several of us were raising the flag saying, ‘I have these problems too’.”* (P11)

> *“Once the disease is identified, once it’s confirmed, what I experienced is that the specialists basically say: ‘Now you’re on your own.’ Because the disease isn’t well known. But, if there were, say, a portal that listed the disease I have, and explained the symptoms and what someone can do, that kind of support would be really helpful.”* (P9)

When only a small number of individuals live with a particular rare condition, the information available to patients about symptoms and disease progression is often very limited. This lack of documentation can be highly stressful for patients and families, leaving them feeling deeply isolated. Future digital solutions should aim to provide trusted and personalized information that helps patients and families understand the condition and anticipate what the future may hold:

> *“The illness I have, the only information I can get is through general internet searches. Because when you ask your neurologist, they say: ‘The illness is known, but the symptoms are not.’ The general practitioner says pretty much the same thing. Very few people have the illness I have. So, you end up alone. There’s no digital support, no help… So, you’re left on your own to find information. We’re kind of in the dark.”* (P10)

> *“I have a disease that is considered very rare. In Quebec, maybe only about 40 people have my specific form, because there are over 100 forms of this disease. When I got my diagnosis, I said: ‘What is this?’ I was told: ‘Look it up online, you’ll find answers.’ It was overwhelming because all the forms* [of the disease] *were mixed together. I found it really difficult. I looked for scientific literature because I have a scientific background, but at some point, I couldn’t understand anything. I’m not a geneticist.”* (P9)

Some participants also suggested that Artificial Intelligence (AI) could be helpful to detect unknown signals from raw patient data and to identify common patterns across individuals:

> *“As we know, with rare diseases, even if we report a slightly unusual side effect from a medication, unless it’s very serious, it won’t be reported in a registry. If I could share access to my symptoms, my medications, and all that, maybe artificial intelligence would be able to see patterns based on the information collected.”* (P10)

#### (ii) Finding specialists or specialized clinics

The need most frequently rated as moderate to high by survey respondents was the ability to identify specialists or clinics with expertise in their specific rare disease. This priority was further reinforced during the focus groups by many participants:

> *“I think the biggest gap in all of this is finding a specialist who truly understands the disease and can provide proper follow-up.”* (P3)

A participant also mentioned the need for greater transparency regarding waiting times to see specialists:

> *“‘I’ll refer you to a neurologist.’ Okay, how long before they call me? ‘I don’t know… maybe in a few months you’ll get a call from a neurology clinic.’ Okay, but where am I in the queue? Am I number 2,000? 400? 60? I don’t understand why there isn’t a system that tells you exactly where you are in line. To me, this could easily be solved with a digital system, like when you call Bell Canada and they tell you that you’re number 7 in line. It’s simple.”* (P7)

#### (iii) Managing healthcare trajectories

Participants expressed many concerns about having to deal with different siloed applications. They expressed a desire for an app that centralizes all functionalities and integrates all aspects of disease management:

> *“It’s about bringing everything together: reminders, the list of medications, the list of doctors with their contact information, ideally a list to track symptoms, a calendar to note appointments… I’m probably forgetting something, but… An information section too. So, providing information about rare diseases. Instead of having 10 different apps, having one that brings it all together.”* (P10)

> *“Having tools like that 25 years ago would have made a big difference… managing all the different appointments. You know, we’d see the doctor maybe once every three weeks, but in between, there were physiotherapy and occupational therapy sessions, so we had quite a few appointments. Three to four times a week in different places. So having something centralized would have been really useful.”* (P12)

Participants emphasized the need to communicate more easily with healthcare professionals online, through teleconsultation, chat or text message for questions about side effects or for follow-up appointments that do not necessarily require physical exams or medical tests:

> *“With doctors in Canada, I don’t have access to Zoom. But, when I went to Central America to get a diagnosis, I used Zoom. Here in Canada, doctors didn’t want to talk via Zoom or WhatsApp. In Mexico, the doctors always communicated with me through WhatsApp and Zoom.”* (P7)

Some participants, especially those in remote areas, also highlighted the challenges of attending in-person appointments and expressed interest in tools that would enable self-management of symptoms, and allow adjustments to care and treatment plans. Many emphasized that healthcare professionals should have real-time access to these tools to facilitate follow-ups and ensure timely care adjustments:

> *“Regarding symptoms, what would be useful would be to manage them ourselves. Like me, I would enter my symptoms and create a database, and my doctor or urologist would have real-time access to it. Plus, I live in a rural area, and I have to call Quebec City or Montreal, and it’s complicated to get appointments.”* (P1)

Similarly, people living with rare diseases often have frequent appointments with various healthcare professionals and must regularly consult specialists located in urban centers, far from their place of residence. In addition, many individuals with rare diseases face physical limitations, making access to teleconsultation or online appointments even more relevant:

> *“One hour to get there, one hour to come back, two hours waiting on an uncomfortable chair, being exposed to infections in a clinic or hospital that worsen my symptoms. And it’s exhausting. All that just to be told: ‘everything looks good, see you in six months.’ With the start of the pandemic, we saw that it was actually possible to do things differently. Of course, a gynecological exam is hard to do virtually, but still, there are so many things that can be done through teleconsultations. It’s so helpful. All the energy I save. I’ve been prioritizing this for five years now, and I avoid clinics and all that like the plague. I haven’t had an infection in five years. And the time I used to lose going to appointments; it could be 5 to 20 hours a week before the pandemic. Before, I couldn’t work more than 10 hours a week. But now, I work almost full-time because I can use my energy for that.”* (P10)

Beyond general care trajectory needs, participants emphasized the importance of having a digital tool specifically designed to support care management during medical emergencies. Across all focus groups, participants described the considerable burden of receiving urgent care from healthcare professionals unfamiliar with their rare condition. Emergency situations were portrayed as highly stressful for patients and families, who expressed fear of inadequate care:

> *“Sometimes, having to explain everything all over again can be long and emotionally draining. That’s how it is for me. Whether it’s in pediatrics, genetics, or elsewhere, I think it would be useful to have a tool, whatever the format, that says: ‘My child has* [name of the syndrome]*.’ But then again, do the people in the emergency room at that moment know about this syndrome? Are they familiar with its characteristics, symptoms, and everything that comes with it?”* (P14)

To address this issue, some participants suggested designing a digital tool that patients could use to inform emergency medical staff about the appropriate care protocol. Such a tool could include a summary of the patient’s health record along with recommended treatments, medications, and specific care procedures. Interestingly, focus groups with patients and families revealed that participants would feel more comfortable using a digital tool to mediate interactions with health professionals rather than providing oral explanations:

> *“It would be helpful to have a summary page with all the relevant information. For example, in my son’s case, children with his syndrome typically don’t have an intellectual disability and aren’t autistic, but my son does have an intellectual disability and is autistic. Sometimes people question that. So yes, the syndrome should be listed, but also the specific characteristics of the person. Because if someone sees the name of the syndrome and already knows about it, they might just glance at it and miss important details.”* (P14)

> *“It’s about having a protocol available when you arrive at the emergency room. One that clearly states what the problem is, what your rare disease is, and what needs to be done. If you arrive in anaphylactic shock, like I did, you say you have* [name of rare disease]*. They respond, ‘What’s that?’ So, I pull out the protocol written and signed by the doctor who diagnosed me. If I arrive at the emergency room unconscious, they have access to some information. They could know what’s going on.”* (P3)

Participants envisioned this tool in various formats, either implemented as an alert embedded within current healthcare systems, such as the Québec Health Record [31], or integrated in their provincial health insurance card (RAMQ card), or another portable device (e.g., a bracelet that could be scanned upon arrival at the emergency room). To be effective and widely adopted, participants stressed that the tool had to be approved by relevant regulatory authorities.

#### (iv) Monitoring adverse effects of treatments or medications, whether severe or not

Across all focus groups, participants expressed strong interest in a digital tool that could help them manage their clinical signs and adverse events following treatment. They also emphasized the importance of accessing reliable information to determine whether these symptoms or side effects are expected or documented for their condition:

> *“I knew something was wrong, but when I searched the internet for my symptoms, nothing came up. If there had been a place I could have written down my symptoms, that would have been definitely helpful.”* (P5)

> *“…to ask my questions. Like a virtual assistant or a database. I use Facebook for that. Otherwise, I will call the neurologist directly because the professionals here aren’t trained, and it’s a bit disjointed.”* (P1)

One participant mentioned how these digital tools could help reduce the burden of managing a rare disease:

> *“I go online to look up my medication, ‘Is what I am experiencing normal, or should I call my pharmacist?’ It* [tool] *would definitely make things easier. Because when you have a rare disease, you spend energy on everything: managing the illness, appointments, going to the pharmacy… When I’m worried, I search online or call my pharmacist because it’s hard to reach doctors. Any tool that could lighten that load would be incredibly helpful.”* (P6)

#### (v) Accessing research related to their rare disease and identifying available clinical trials

Participants also mentioned additional needs related to clinical research. Several expressed a desire to be better informed about ongoing studies, particularly to know whether any research teams are currently working on their specific condition. They also suggested greater patients’ involvement in research, including collecting information about their signs and symptoms, with data being reusable for future research:

> *“I really believe that when someone receives a diagnosis, they should be involved in research. They should be registered in research. Right now, we’re not trying to address the root problem. We’re just treating symptoms here and there. Like, ‘Oh, you have inflammation’. But what’s causing the inflammation? Instead of addressing the source, we’re just patching things up. So, patients diagnosed with rare diseases should be automatically registered in research, so we can find solutions to the root causes, not just the small symptoms.”* (P4)

> *“There should be some kind of database where people with the disease could go and say, ‘I have this symptom’ and researchers could look into it. It could help generate research based on data that isn’t yet conclusive but could become so.”* (P11)

One participant even expressed interest in accessing simplified clinical and scientific conferences on their rare disease, featuring specialists and researchers in the field. For example, in a Q&A format:

> *“From time to time, we have access to clinical conferences. For example, a urologist comes to explain certain things to us, but in simplified terms. It’s just for patients, and that’s really appreciated. Because otherwise, we don’t get that kind of information. It’s not during my appointment with the urologist… I get exactly 10 minutes. And if there’s a delay, she can’t answer my questions.”* (P11)

### Digital tools to support self-management of daily life activities

Survey results indicated that 69.1% of respondents showed moderate to high interest in digital tools designed to support daily life activities (e.g., adapting diet or environment), 73.8% in tools for psychological well-being, 81.2% in tools to locate social services or assistance programs, and 71.8% in platforms for connecting with others who share similar experiences.

During focus groups, some participants highlighted the challenge of bridging the gap between their diagnosis and the non-medical services available that could support daily life:

> *“I wonder if there could be a link between the diagnosis and the CLSC* [refers to a local community services center in Quebec]*? Because, honestly, we don’t know what services they offer. When we go there to try to access them, it’s like looking for form A38 in the madhouse! You’re sent from one department to another. Just getting a parking permit is complicated. These are the kinds of things that make the process long and difficult.”* (P13)

Moreover, as with other chronic diseases, they expect greater access to telehealth services when attending in-person appointments is difficult, whether due to remote living, disabilities, or external factors such as pandemics:

> *“During the pandemic, my son wasn’t in school. He was attending virtual classes. So, during that time, he didn’t have access to his physiotherapist, his occupational therapy, or his speech therapy, but his illness didn’t stop. So, what was I supposed to do? I had to pivot and search online. I found nothing. What exercises are appropriate for this type of child? At the very least, a reference that says, like P13 mentioned, how to get a disabled parking permit, what resources are available? A single website… Maybe a mega-site. It’s a bit of a dream, but right now, there’s nothing.”* (P14)

One participant also expressed the need for digital tools to increase autonomy at home, particularly to automating daily tasks and easing life with visual impairment, for example:

> *“Technological tools to help me go about my daily activities, yes, I need them. Some have been put in place, especially for my visual impairment. But I think there’s still room for improvement. I have a door unlocking system, but I have needs in many other areas: for fall prevention, for lighting in the house, for accessing objects that are too high, for lowering blinds, for many things… Basically, home automation. Yes, technology could be integrated into our homes to help us maintain our autonomy. Technology will allow us to preserve it.”* (P8)

Several focus group participants emphasized the need for peer-support communities where they can exchange with others who understand the disease journey, access appropriate resources, receive support and advice, as well as validate symptoms that may be unfamiliar to healthcare providers. Many currently use social media to engage with these communities and share experiences with other rare disease patients and families:

> *“Sometimes, when you’ve just been diagnosed and you’re on the verge of a crisis, and things aren’t going well, it can really help to talk to a human being* [other participants nod in agreement]*. So, there’s that human aspect. I was lucky, I got to talk to someone who had been living with this disease for a while. She was able to give me advice. She’s not a doctor, but… So yes, that human connection can be really valuable. Maybe starting with a chat message or an online appointment. Even that can make a big difference when you don’t really know what’s happening to you.”* (P5)

> *“Sometimes I find it a bit comforting. Like at one point, I was really experiencing ‘brain fog’, and I found it so hard to live with. So, I brought it up in a Facebook group* [a group related to her rare disease]*. I asked, ‘Is anyone else experiencing this? Because it’s not mentioned in the literature.’ Same thing with cognitive issues – they’re not addressed in the scientific literature. And at one point, I started panicking. I thought, ‘What’s going on? I’m not supposed to have cognitive issues.’ I was thinking, ‘My goodness’, on Facebook, when we talk, we mention so many symptoms that aren’t necessarily listed anywhere.”* (P9)

> *“In those Facebook groups, there’s as much junk as there is validation for what I’m going through. You know, I tell myself, ‘It’s not just in my head.’ At one point, my hands were clenched. I had cramps in my hands; they turned into claws. That’s not in the literature, because in the literature, they focus more on genetic research to find a treatment. But in the meantime, we’re the ones living with these problems.”* (P11)

> *“Sometimes it’s also about practical tips. Like, I had this issue: I dislocated my hip. How can I put it back? But if you call your doctor, they’ll say, ‘Come on, it’s not possible to dislocate your hip, that’s really rare.’ You know, they don’t understand the reality. So, it’s kind of all that.”* (P10)

However, many also noted the potential risks of relying on such communities for disease management. They expressed concerns about the accuracy of shared information and emphasized the need for reliable, validated resources on their rare disease and its management. Participants proposed a moderated platform where posts are monitored, sources are clearly identified, and trusted individuals provide accurate details about the rare disease:

> *“People with my disease tend to rely on Facebook groups a lot. But with Facebook groups, you have to take things with a grain of salt. Some people ask a question, and the answers make no sense. There are those ‘know-it-alls’.”* (P8)

> *“In Facebook groups, some people start acting like amateur doctors: ‘I got this number in my test results, what do you all think?’ So, it’s like we’re trying to self-analyze things that should really be interpreted by specialists. I think it leads to a kind of drift. Like for my hypochondriac mother-in-law, one little number slightly above normal, and suddenly she thinks she has cancer. So, we really need to be careful.”* (P11)

### Improving care coordination through real-time and centralized access to information

Survey results showed that 79.0% of respondents expressed moderate to high interest in exchanging information directly with their healthcare professionals, reflecting a strong need for interactive and accessible communication (**Table 3**). Additionally, 74.5% indicated interest in recording information and sending real-time alerts to their providers.

**Table 3.** Survey respondents’ interest in real-time data sharing with their medical team (n=149)

|  | n (%) |  |
| --- | --- | --- |
|  | Moderate to high interest | Little to no interest |
| To exchange information with your medical team. | 119 (79.0) | 30 (20.1) |
| To record information and send real-time alerts to your medical team. | 111 (74.5) | 38 (25.5) |

Information sharing with healthcare providers and data centralization emerged as key themes in the focus groups. Several participants reported having to maintain an updated medical file at home, including health details, exam and test results, as well as appointments with various professionals, to ensure proper coordination and continuity of care. However, some noted that this responsibility resulted in a considerable cognitive burden, adding to the challenges already posed by their illness:

> *“We need to keep meticulous records at all times. Everything, absolutely everything. As users, we have to request, archive, and carry all this information with us to every specialist we see. Having it in a digital format would make a huge difference. That kind of access would significantly simplify things and lift that responsibility off our shoulders. […] It would be much easier to have something highly centralized and digitized*.

> *Something that could be placed at the core of the person’s condition and shared with everyone involved in their care. That way, access would be much simpler. Because right now, there’s clearly a lot of repetition. What’s really needed is the implementation of a program or software that coordinates our medical file, so that all professionals working around us can access everything.”* (P8)

> *“Information sharing between us* [patients] *and the doctors, and vice versa, is something that’s been available for a long time in the United States. There are portals where patients can send questions to their doctor or clinical team, upload documents, and sometimes even track symptoms or medications. The clinical team can access that information, and the exchange goes both ways. Here, it takes about a month before we can access our own data.”* (P10)

### Priorities for digital health in rare diseases

More than half of survey respondents (53.7%) identified medical care management as the top priority for implementing digital solutions in Quebec (**Table 4**). Meanwhile, 26.2% prioritized access to useful information about their rare disease, and 20.1% focused on digital tools to support day-to-day management.

**Table 4.** Survey respondents’ priorities for implementing digital tools for rare diseases (n=149)

| **Priority** | **n (%)** |
| --- | --- |
| To improve the medical management of my rare disease or the rare disease of the person I am caring for. | 80 (53.7) |
| To access useful information about my rare disease or the rare disease of the person I am caring for at different stages of my/their medical journey. | 39 (26.2) |
| To learn to better live with my rare disease or the rare disease of the person I am caring for on a day-to-day basis. | 30 (20.1) |

Focus groups priorities aligned closely with survey findings. Half of the participants (7/14) identified patient information centralization as the top priority for implementing digital tools in Quebec. Patients and caregivers expressed a strong need for a solution that enables collecting and sharing information with physicians and all healthcare professionals involved in their care.Access to comprehensive, fast (ideally real time), and user-friendly patient data was seen as essential for improving care coordination and management.

Other priorities included developing a digital assistant to mediate emergency interactions with medical staff (3/14), improving access to teleconsultation or telemedicine services (2/14), creating a peer-support community (1/14), and enhancing education and awareness among healthcare professionals about rare diseases (1/14).

### Concerns about the use of digital tools in rare diseases

Overall, survey respondents did not express significant concern about most potential issues related to digital tools (**Table 5**). However, nearly half reported moderate to high concern about two issues: protecting personal information and losing human interaction.

**Table 5.** Survey respondents’ concerns related to the use of digital tools for the management of rare diseases (n=149)

|  | n (%) |  |
| --- | --- | --- |
|  | Little or no concerns | Moderate to high concerns |
| Protection of personal information or privacy | 75 (50.3) | 74 (49.7) |
| Feeling of losing human contact | 78 (52.3) | 71 (47.7) |
| Potential costs associated with using digital services and tools | 92 (61.7) | 57 (38.3) |
| Difficulty using digital services or tools due to a disability, when applicable | 119 (79.9) | 30 (20.1) |
| General health knowledge | 135 (90.6) | 14 (9.4) |
| Computer skills | 142 (95.3) | 7 (4.7) |
| Ability to access the internet | 143 (96.0) | 6 (4.0) |
| Information-seeking skills | 146 (98.0) | 3 (2.0) |

These concerns were also raised by some participants during the focus groups. One participant specifically expressed worries about data security when using digital tools:

> *“It’s great to have digital access, but we also need to ensure data integrity. That it* [the data] *isn’t tampered with, and that it remains protected from unauthorized access. These processes are important. Yes, it’s good to democratize access and have digital tools, but we also need to think about security.”* (P12)

Another participant spoke about the loss of human connection associated with digital tools, describing them as sometimes cold and impersonal:

> *“It’s hard sometimes, you know, because I’m not feeling well, it’s a lot. That’s why computers… there are no emotions* [emphasizing her preference for in-person contact over internet-based communication]*. Maybe it’s easier as a tool, but still, yes, talking to someone in person… so we can talk when things aren’t going well.”* (P2)

Other participants raised additional concerns about the implementation of digital tools for managing rare diseases, issues not addressed in the survey. One participant pointed to a broader systemic challenge within Quebec’s healthcare infrastructure, such as lack of funding and limited resources, expressing skepticism about the system’s ability to implement such tools:

> *With everything we know about the healthcare system, budget cuts and all, it’s already very difficult to implement digital systems. So now I’m thinking, how are they going to implement new systems? It takes funding, it takes political will, it’s a heavy process. I mean, I don’t want to discourage your study, but everything people have said* [referring to what participants said in the focus group]*, it would be amazing to have all that. But how are we actually going to make it happen? That’s what concerns me.”* (P6)

## Discussion

This study aimed to explore the needs and priorities of rare disease communities regarding health digital tools. Both survey and focus groups participants reported challenges similar to those identified in previous studies and reports, contributing to the definition of shared priorities [4,28,32–34].

Based on our findings, the primary benefit expected from digitalization is improved **information exchange between healthcare professionals and patients/caregivers**. Participants expressed concerns about siloed data and emphasized the need for a more integrated health information system. These concerns echo the long-standing call for greater interoperability, a demand voiced for years by healthcare professionals and IT system developers. A patient-centric information system is considered a top priority for rare disease communities, given the chronic and complex nature of these conditions, which require high levels of care coordination. Such a system should not only allow patients to access data entered by professionals but also enable them to contribute by collecting patient-reported information and outcomes. Participants expressed a strong desire to act as data providers. Therefore, the priority is to break down silos, establish a pan-institutional model, and develop coherent policies and IT-frameworks that reduce fragmentation and support an inclusive vision of patient care. Most participants were enthusiastic about creating user-friendly communication tools to connect patients with healthcare professionals, suggesting solutions ranging from secure platforms to integrated messaging features.

Patients and caregivers advocated for **telemedicine** as an alternative to in-person visits. Given the very low prevalence of each rare disease, only a few medical experts possess extensive knowledge of specific rare diseases. As a result, some patients must travel long distances for consultations. All telemedicine modalities were mentioned by at least one participant. Teleconsultation was mentioned as a way to improve access to specialized care, while remote health monitoring was seen as a means to support more personalized, real-time care. Building on this idea, several focus group participants emphasized that healthcare professionals should have real-time access to patient data to enable timely adjustments to care and treatments. However, such an approach raises concerns about potential “alert fatigue” among clinicians managing large patient populations. To address this challenge, implementing AI-driven alert triage algorithms will be essential to minimize alert fatigue while supporting real-time, precision-oriented care [35]. Beyond medical needs, nearly 70% of survey participants expressed moderate to high interest in digital solutions to help overcome **daily life** challenges. Social media networks were also mentioned as essential for accessing peer-support communities and sharing experiences. Several focus group participants viewed these platforms as a way to mitigate feelings of isolation. A similar point was raised by Chang et al. [28]: “*Having a community who has been through similar experiences helps to mitigate this isolation. These community members truly understand the highs and lows of managing a rare disease, and can offer the emotional support and motivation to keep going*.”. Given the growing popularity of social media, with 5.66 billion users worldwide as of October 2025 [36], and the isolation experienced by many rare disease patients, it is predictable that the number of patients and families using these platforms will continue to rise globally. In line with other authors [37], we argue that community-focused digital tools are central in the rare disease domain and that healthcare professionals should pay close attention to patient discussions on social media. Furthermore, knowledgeable health professionals could play an active role in ensuring that circulating information is accurate and reliable.

In addition, focus group participants expressed a strong interest in **modern digital tools using state-of-the art technologies**. Rare disease patients are eager for user-friendly applications that support multiple languages and adapt to varying levels of health literacy. When seeking disease-related information or scheduling specialist appointments, participants often compare existing tools for rare disease patients with commercial applications developed by private companies. Their conclusion was consistent: future dedicated tools should integrate the usability and design standards of the apps they are already use in daily life. One participant noted that, although relevant disease-related information may be available on websites such as Orphanet (orpha.net), it often remains difficult to access due to poor usability. Based on focus group discussions, a major barrier to adopting digital health solutions is that existing services fail to meet consumer expectations. This observation aligns the growing popularity of generative AI (GenAI) tools in the general population and the increasing role of LLMs and AI in digital health [17,38–40]. However, evaluation protocols for these tools are not standardized, and GenAI-based solutions are constantly evolving. Consequently, despite the abundance of literature, providing clear scientific evidence on their benefits and limitations remains challenging [41]. Recent studies assessing patient perceptions of GenAI responses indicate that most patients (e.g., 78% in Ayers et al. [17]) preferred chatbot answers over physician responses, describing them as “more empathic”. Yet, the issue of clinician oversight remains [42]. Following recommendations from the American Medical Associations [43], we argue that engagement with AI in rare disease care should be limited to tools that meet rigorous standards for advancing health equity, prioritizing patient safety, and minimizing risks. Furthermore, patients highlighted the potential of AI to generate new knowledge from patient-generated data, such as identifying patterns in datasets of self-reported symptoms and medical events.

Regarding the **impact of digitalization**, the proportion of participants who expressed concerns about data privacy was roughly similar to that reported in surveys focusing on non-rare disease populations, such as pregnant women [24]. In our study, while 95% of respondents are fully familiar with digital tools, only half of them expressed concerns about data privacy. It appears that rare disease patients expect more benefits from digital tools, and even data sharing, than the average citizen. Several studies involving patients and the general public have reported concerns about data security and confidentiality in mobile health applications or in data sharing in general [44–47]. Compared to the general population, patients with rare diseases seem more willing to share their data and tend to be more motivated by the prospect of improving care and research [45,48]. Despite this, several studies indicate that they remain sensitive, sometimes even more so, to issues of confidentiality and privacy protection [23].

Another noteworthy finding from our study is that **digital tools may alter the respective roles of rare disease patients and physicians**. Half of the participants anticipated that digital tools could lead to a loss of human contact. This may affect physicians’ roles, especially since many participants reported challenges in their relationship with their doctors. Several participants highlighted the limited knowledge of rare diseases among health professionals, as well as difficulties in being taken seriously and even listened to by medical staff. They described these challenging interpersonal dynamics as a major barrier within their care pathway. The so-called “medical gaslighting” [49], a phenomenon in which healthcare professionals dismiss or minimize patients’ symptoms without proper medical evaluation, often due to a lack of awareness or understanding of rare diseases, is a real concern for several patients and families in our study [50,51]. The combination of isolation and experiences of gaslighting among individuals with rare diseases makes seeking online information and adopting digital tools more appealing. Traditionally, medical decisions regarding diagnosis and treatment are made by clinicians, or clinicians provide explanations to patients to achieve shared decision-making. In our study, some participants expressed a desire for patient-centric digital solutions that would enable them to participate in decision-making. This demand for greater patient empowerment was most pronounced in two highly stressful situations: first, delays in diagnosis; second, emergency care. One participant shared a vision that resonates broadly within rare disease communities [52,53]:

*“A Quebec tool that you could bring to your doctor and say, ‘I looked this up, what do you think?’ We are the patients; we are the ones who know the most about our condition”.* Notably, our survey revealed that 22.5% of our participants experienced a diagnostic odyssey, receiving a diagnosis after five years. Among them, five participants waited 20 years, confirming the need to prioritize digital solutions that can help reduce time-to-diagnostic. In emergency situations, rare disease patients and caregivers expressed interest in digital tools to facilitate communication with emergency medical staff. An app acting as a knowledgeable assistant was mentioned in every focus group, highlighting its potential as a priority for reducing anxiety among families and minimizing the risk of medical errors.

Our findings suggest **that joint international efforts are required to address patients’ expectations**. Some participants expressed concerns about cuts to health budgets that could slow the development of digital tools in many countries. Moreover, the existence of a myriad of rare diseases, with different symptoms, genetic profiles, progression and associated disabilities, will require joint interdisciplinary collaboration, even partnerships between the private and public sectors to meet patients’ needs. For some ultra-rare diseases, there are only a few experts worldwide, and digital tools offer a way to transfer their knowledge into applications, enabling patients living far from specialized care facilities to benefit from their expertise and receive safer care. Similar obstacles exist in developing countries, where healthcare challenges are closely intertwined with economic development and uneven resource allocation. Digital solutions could help increase equity in addressing these challenges and support the WHO’s priority of reducing the global burden of rare diseases [13].

### Study limitations and strengths

This study has some limitations. First, although the survey was distributed through Quebec rare disease associations, 22 survey respondents were from other Canadian provinces. Differences in healthcare systems and priorities may limit comparability, but their responses were still included to capture general needs for digital tools. All participants in our study were volunteers, and no probabilistic sampling was used, which may affect representativeness. We also lack data on respondents’ location, relying instead on travel time to the nearest rare disease specialized facility. As a result, we could not assess urban-rural differences or geographical representativeness, though focus groups revealed specific needs related to remote living. The sample was predominantly female and highly educated, which may influence perspectives on digital tools compared to the broader population. Finally, the study focused on patients and caregivers; clinicians’ views on digital solutions were not assessed, which could have provided additional insights.

Besides those limitations, a major strength was the active involvement of the rare disease community from the outset. The largest organization representing individuals with rare diseases in Quebec (RQMO) contributed to questionnaire design, survey distribution, and participants recruitment. Two patient-partners helped refine and clarify the survey questions and discussion guide, and 15 of 149 survey respondents volunteered for focus groups, demonstrating strong engagement.

## Conclusion

In conclusion, our findings align with previous studies [28] and underscore the central role of digital solutions for people living with rare diseases. Several priorities emerged from the challenges reported by patients and caregivers in Quebec, including the need for a system that centralizes health data, supports patient-generated information, and enables exchanges with healthcare professionals. Participants also expressed strong demand for telehealth services to reduce in-person visits and simplify daily life, as well as tools that place patients at the center of care throughout the diagnostic process and in emergency situations. Building on these insights, digital solutions should rely on modern technologies, integrate data across the health system, support telehealth, incorporate AI algorithms, and offer functionalities to manage the most stressful situations. Such tools, if trustworthy, could significantly reduce the burden of living with a rare disease. While technological, legal, political, and resource-related challenges remain, these findings help identify key targets for future interventions.

## Availability of data and materials

The survey questionnaire and the discussion guide used for the focus groups are available as Supporting information files. All data generated or analyzed during this study are included in this published article.

## Competing interests

The authors have no competing interests to declare.

## Funding

This study was conducted as part of the activities of the *Chaire MEIE du numérique en santé*, funded by the *Ministère de l’Économie, de l’Innovation et de l’Énergie* (MEIE) and the *Université de Sherbrooke*. Additional funding was provided by the *Groupe de recherche interdisciplinaire en informatique de la santé* (GRIIS) and the PR[AI]RIE Institute.

## Authors’ contributions

AB, CK, and JFE were responsible for project administration and financial resource management. AB, CK, RD, and JFE contributed to the study’s conceptualization, methodology development, analysis of the results, and critical revision of the manuscript. AB and RD drafted the initial version of the manuscript. RD was responsible for participant recruitment, data collection and thematic data coding using qualitative analysis software.

## Acknowledgements

The authors would like to thank all study participants for their time and valuable contributions. The team also wishes to acknowledge Raphaé Macchabée (RM), an undergraduate student in pharmacology who completed an internship with our team. RM contributed to organizing the focus groups, drafting the discussion guide, and served as a technical assistant during the sessions.

## Supporting information

**S1 Text.** Survey questionnaire.

**S2 Text.** Discussion guide.

## Notes

### Competing Interest Statement

The authors have declared no competing interest.

### Author Declarations

Ethics approval was obtained from the Educational and Social Sciences Research Ethics Committee of the University of Sherbrooke, Québec, Canada (approval number: 2023-4224). Informed consent was obtained online from all participants prior to their involvement in the study. Participants’ contact information was stored in a secure online file accessible only by authorized members of the research team. Additionally, the qualitative data, such as the discussion transcripts, were stripped of all identifying information to preserve participant confidentiality.

